# Dysregulated Immune Proteins in Plasma in the UK Biobank Predict Multiple Myeloma 12 years Before Clinical Diagnosis

**DOI:** 10.1101/2025.02.04.25321690

**Authors:** Josh Fieggen, Anshul Thakur, Christopher C Butler, Karthik Ramasamy, Anjan Thakurta, David A. Clifton, Lei Clifton

## Abstract

High-throughput proteomics has emerged as a potentially rich data source to improve capacity to forecast disease. This study explores the utility of plasma proteomics for identifying novel predictors of Multiple myeloma (MM), combining machine learning with statistical approaches. Utilising data from the UK Biobank, including proteomic profiles of over 50k participants, we applied an “extreme gradient boosting” (XGBoost) algorithm with SHapley Additive exPlanation (SHAP) feature-importance measures to identify key proteomic biomarkers to predict onset of MM. At least seven of the top 10 identified proteins are related to immune function and activation of lymphoid cells; two are validated MM targets with approved therapies. The top 10 proteins along with key clinical predictors were further analysed using Cox proportional hazards models to assess their contribution to incident MM risk. 10 proteomic biomarkers ranked by SHAP value substantially outperformed traditional clinical predictors. This superior performance was maintained over the 12-year follow-up period, demonstrating the predictive ability of these proteomic biomarkers for early detection of MM. The demonstration of the dysregulated expression of proteins in serum from healthy individuals, if confirmed in prospective cohorts and independent datasets, could lead to novel approaches to screening for MM and precursor conditions.

## Background

Multiple myeloma (myeloma) represents a significant clinical challenge due to its symptom burden at diagnosis, often due to delayed presentations^1^. There are few specific risk factors for myeloma and diagnosis is often only made following complications such as anaemia, bone lesions, renal failure, and immune dysregulation.^2^ While myeloma remains incurable, early diagnosis is critical to improving outcomes.^3^

Proteomics has emerged as a pivotal tool in cancer research, offering insights into the molecular basis of various malignancies.^4^ Early myeloma and its pre-cursor disease states have high quantity of immunoglobin paraprotein in blood as a key marker of disease.^2^ In addition, levels of albumin, b2 microglobulin (B2M), and lactate dehydrogenase are assessed to risk-stratify patients.^5^ However, a proteomics-based diagnostic has not been developed for myeloma. It is also plausible that plasma from healthy individuals may contain proteins from organs and or cells that serve as biomarkers of physiological dysregulation that precedes the onset of disease. Availability of Olink plasma proteomic data^6^ of 2932 unique proteins from 54219 healthy participants with a long-term clinical follow up in the UK Biobank^7^ (UKB) allowed us to explore the latter.

## Abbreviated Methods

Our study population includes participants with baseline plasma proteomics data, excluding prevalent myeloma cases (those with existing diagnoses at baseline). Our disease outcome is defined as incident myeloma cases (diagnoses of myeloma after the baseline date) ascertained via linked cancer registry, death registry and in-patient hospital records. To identify the top 10 proteins predictive of myeloma we employ a machine learning based feature selection pipeline (Figure 1a) using an “extreme gradient boosting” (XGBoost) algorithm with a Cox loss function and SHapley Additive exPlanations^8^ (SHAP). These were then used with, and in comparison to, the best available clinical variables known to predict myeloma in the general population^9^ (Supplemental Method 1). To do this, three Cox models (one using clinical variables, one with proteomic biomarkers, and a third combined model incorporating both) were developed in 80% of the data and tested on the remaining 20%, with performance assessed using time-dependent receiver operating characteristic curves and concordance indexes. Detailed statistical methods are described in Supplemental Method 2.

**Figure 1:**
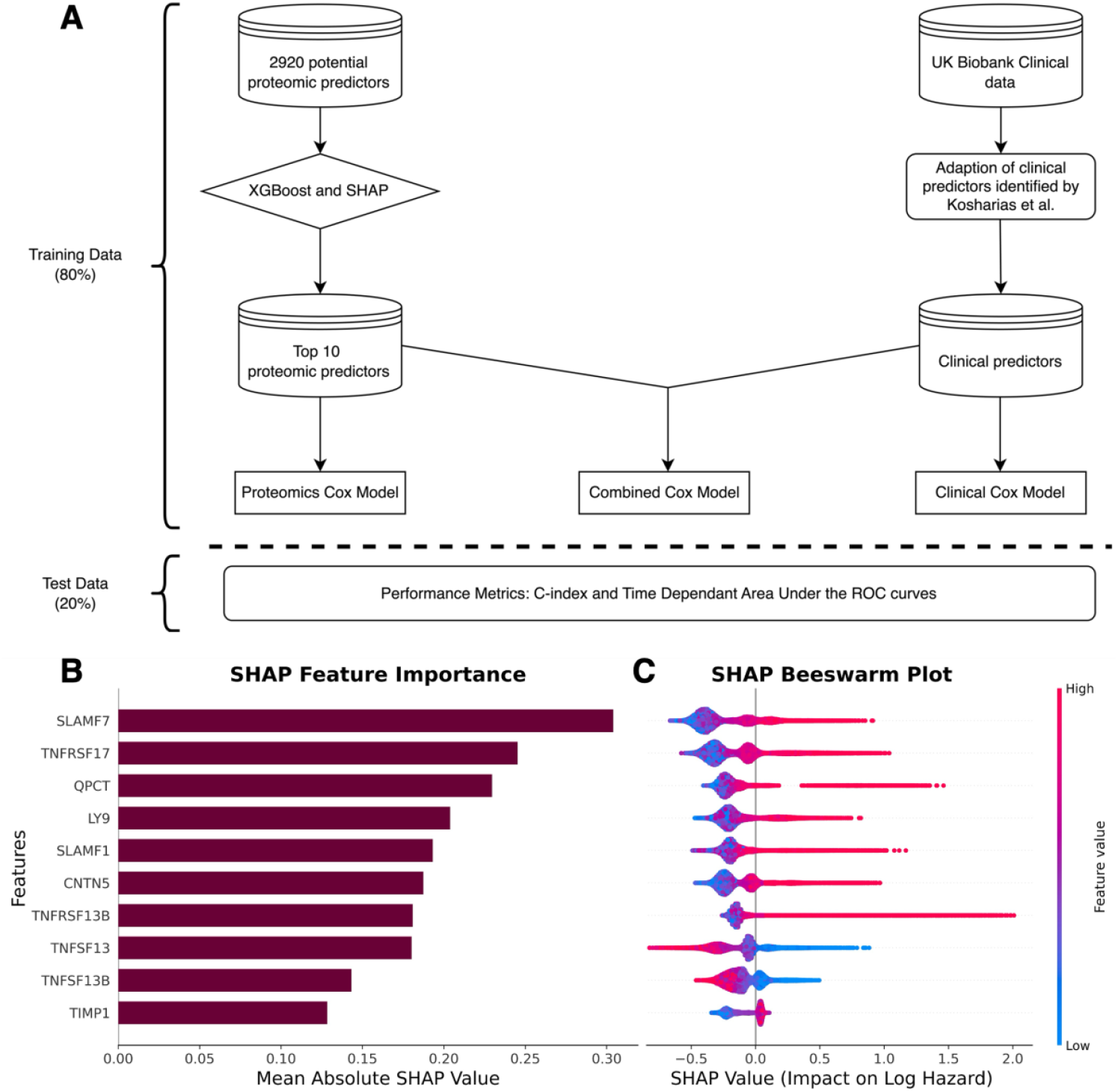
Model development pipeline and top proteomic machine learning model features. **Panel A** outlines the pipeline used to predict myeloma by integrating proteomic and clinical data from the UK Biobank. Starting with 2920 potential proteomic predictors, a tree-based XGBoost algorithm combined with SHAP values was employed to rank and identify the top 10 predictors. These were then used to develop a **proteomics Cox model**. Clinical predictors including age, sex, symptoms, and haematological parameters were used to develop a **clinical Cox model**. Finally, the top proteomic and clinical predictors were combined to create a **combined Cox model**. All models were evaluated on test dataset, with performance assessed using the concordance index (C-index) and time-dependent area under the receiver operating characteristic curve. This pipeline demonstrates the combination of advanced machine learning with traditional modelling to enhance the prediction of myeloma. **Panel B** displays a bar plot of the mean absolute SHAP values for the top 10 features. In the context of a model with a Cox-loss function, a SHAP value represents the marginal contribution of each feature to the log-relative hazard (i.e. risk score) from baseline for an individual. This panel provides a summary of the average of all individuals contributions to the model’s predictions. The features are ranked, with higher values indicating greater importance in influencing the model’s output, providing a comparison of which proteomic markers are most critical in ranking myeloma hazard. **Panel C** displays a scatter (“beeswarm”) plot where each dot represents an individual data point in the dataset. The points are distributed horizontally along the x-axis according to their SHAP value. Where there is a high density of similar SHAP values, points are stacked vertically. The colour of the dots reflects the feature value, with red indicating high feature values and blue indicating low feature values. The plot provides a granular view of how each feature contributes to the prediction at an individual level. It shows the distribution of SHAP values for each feature, revealing how consistently (or inconsistently) a feature affects the model’s output across different data points. Features with a wide range of SHAP values indicate a strong but varied impact on the model’s predictions, while a narrow range suggests a more uniform influence.

## Results and Discussion

Incident myeloma was diagnosed in 174 (0.3%) of the cohort participants (Supplemental Table 1) with a median time to diagnosis of 7.2 years and whole cohort median follow up of 13.2 years. Participants diagnosed with myeloma were older and more likely to be male. Further baseline clinical characteristics are shown in Supplemental Table 1.

The top 10 of the 2920 features from the optimised XGBoost model, as ranked by mean absolute SHAP value, are demonstrated in Figure 1b, and Supplemental Table 2 describes the function^10^, location^11^, single cell expression^11^, and role in myeloma therapeutics^12^ of these proteins. It is notable that at least seven of these proteins have known biological function in lymphoid cells. This includes three signalling lymphocytic activation molecule (SLAM) family receptors, multiple of which are either current or potential targets for anti-myeloma immunotherapies.^13^ Also identified by the algorithm are the interacting ligands and receptors B-cell activating factor (BAFF), a proliferation-inducing ligand (APRIL), B-cell maturation antigen (BCMA), and transmembrane activator and calcium modulating ligand interactor (TACI) which have known relevance to myeloma pathophysiology.^14,15^ While QPCT and CNTN5 are not currently known to have any clear function related to B-cell biology or myeloma development, both have been noted to be upregulated in plasma cells from some myeloma patients at the single cell level.^16,17^ TIMP1 is a non-specific metalloprotease inhibitor involved in innate immunity.

The individual SHAP values (Figure 1c) show the marginal contribution of each protein to the log-relative hazard (i.e. risk score) from baseline for individuals. The top seven proteins found show a positive association where high relative protein concentrations are associated with higher predicted risk of future myeloma. Curiously, APRIL/TNFSF13 and BAFF/TNFSF13B show the opposite effect where higher relative concentrations are associated with lower risk scores. This appears in contrast with previous literature that suggests APRIL and BAFF are potential markers of myeloma disease activity^18^, however given that these proteins are involved in normal B-cell functioning this may highlight the complex role in the immune dysregulation that precedes the clonal proliferation of malignant plasma cells. When SHAP plots were used to explore interactions (Supplemental Figure 1), there was a clearer interaction pattern identified between TACI and APRIL than between TACI and BAFF despite both these ligands being known to bind TACI.

The distributions of the relative concentrations of the top protein predictors stratified by incident myeloma status are shown in Figure 2a. We use these proteins to construct our first cox model (red, Figure 2b), in which multiple proteomics markers were statistically significant predictors of myeloma. Notably, SLAMF7, TNFRSF17 (BCMA), QPCT, SLAMF1, and CNTN5 were associated with statistically significant higher hazard ratios. Conversely, BAFF (TNFSF13B) had a protective effect in keeping with the SHAP findings. In the clinical model (black, Figure 2b) higher age, male sex, and lower haemoglobin were associated with higher risk. In the combined model (grey, Figure 2b), age remained a significant predictor, while notably sex lost statistical significance, potentially suggesting variance clinically attributable to sex is captured by the proteomics features. The proteomic markers remained significant and almost identical in magnitude to the proteomics model, underscoring their robust association with disease. In a sensitivity analysis excluding cases diagnosed within 5 years (Supplemental Figure 2), the noted proteomic associations largely persisted.

**Figure 2:**
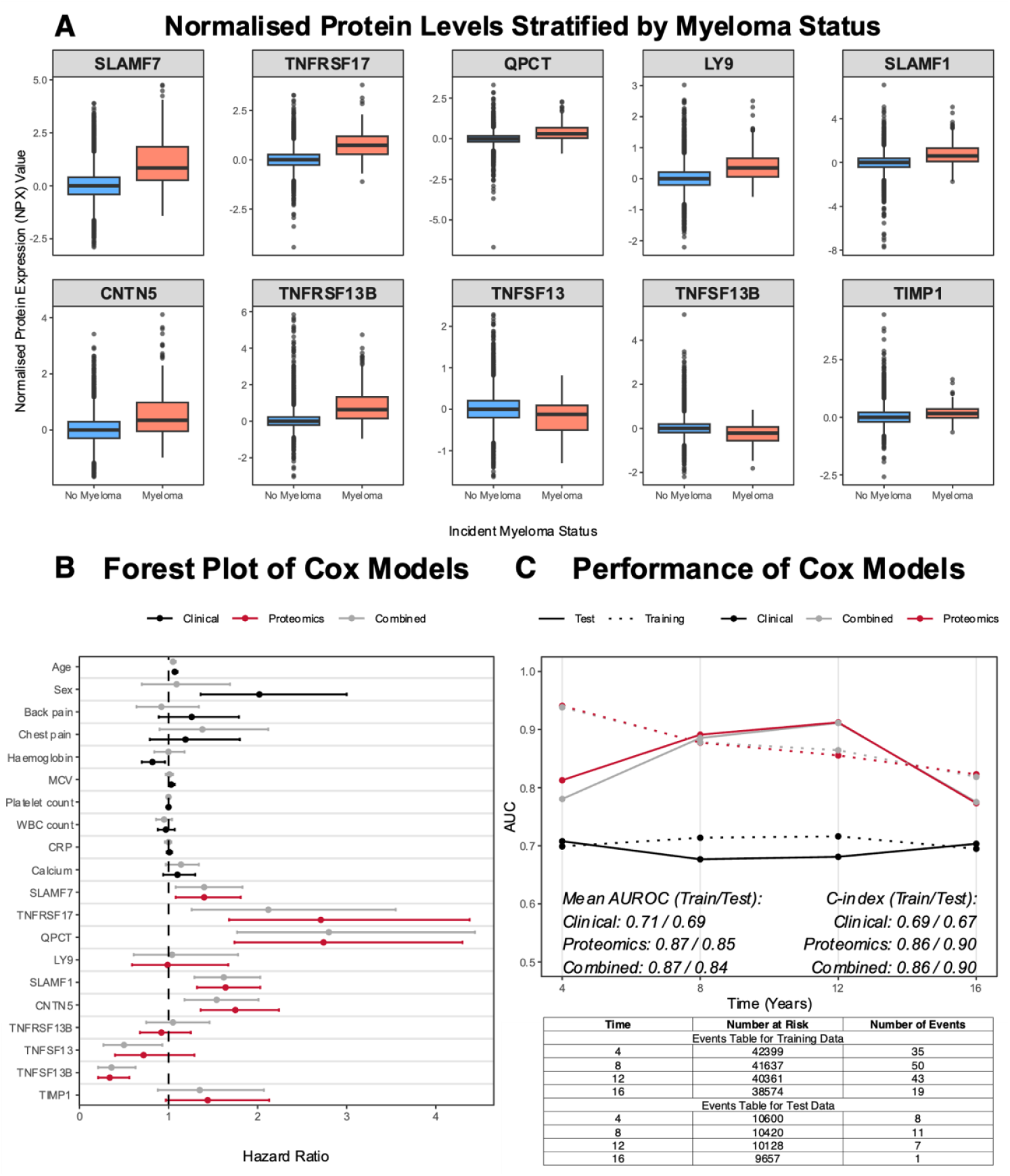
Associations between proteomic and clinical features for incident Multiple myeloma. **Panel A** displays box plots of the Normalized Protein expression (NPX) values of each of the top 10 proteomic features at baseline (enrolment into UKB) values stratified by incident myeloma status. The plots are arranged in order of SHAP importance. Box plot lines represent median NPX value, edges 1th and 3rdth quartiles and whiskers 1.5 times interquartile range with dots as outliers outside this range. **Panel B** shows a forest plot of the Hazard Ratios estimated from the three cox models developed; proteomics in red, clinical, in black, and combined in grey. Hazard ratios point estimates are represented by the dot and 95% confidence intervals are represented by the whiskers. **Panel C** is a comparison of the performance of the three Cox models on the training and held-out test datasets. The line plot displays the time-dependent area under the receiver operator characteristic curves (AUCs) for each of the three models at years 4, 8, 12, and 16 since enrolment in UKB with the mean AUCs and overall C-indexes in the training and test data summarised below. The tables below present the number of participants in the training and test datasets at risk at years 4, 8, 12, and 16 respectively as well as the number events (myeloma diagnoses) occurring between years 0-4, 4-8, 8-12, and 12-16 respectively.

The clinical model had the lowest performance on both the training and test data with C-indexes of 0.69. In contrast, the proteomics and combined models performed very similarly, substantially outperforming the clinical model. Both had C-indexes of 0.86 and 0.90 in the training and test data. Model performance improved in the test data for both the proteomics and combined models at each 4-year time interval until 12 years of follow up. These results suggest plasma may contain biomarkers that long precede disease defining events.

This analysis shows that a hypothesis free and data-driven approach may capture patterns that are reflecting biological B-cell dysregulation that precedes the onset of clinical disease in myeloma. In the context of recent literature potentially supporting population monoclonal gammopathy of undetermined significance (MGUS) screening^19^, better understanding of the mechanisms that lead to progression to myeloma is increasingly important. An important limitation of this study in this respect is the inability to comprehensively describe participant MGUS status or baseline paraprotein concentrations. To attempt to understand what impact this may make, we re-fitted our Cox models excluding all prevalent and incident cases of MGUS (Supplemental Figure 3). This analysis showed that proteomic associations were largely unaffected by the removal of all MGUS cases. Given it has recently been shown that MGUS detected via screening and incidental finding have similar progression risk^19^, this finding gives us more confidence that the identified proteomic associations are important independent of underlying MGUS status.

The increasing attention to MGUS highlights the need for further research to understand how these markers change dynamically as individuals move from a healthy baseline through various precursor states and into clinical disease. In addition, this work should be further developed to explore whether predictive performance can be maintained with fewer proteins and including protein interactions identified by XGBoost, as well as exploring interactions with more commonly measured clinical markers such as total protein and albumin. Orthogonal biological approaches also need to be considered to identify and understand the source and biological implications of the proteins identified. Finally, in future it will be key to distinguish between and optimise models for time-windows relevant to specific clinical questions.

## Acknowledgements

This research has been conducted using the UK Biobank Resource under Application Number 83801. We thank the participants of the UK Biobank study without whom this research would not have been possible. Computing of this study used the Oxford Biomedical Research Computing (BMRC) facility, a joint development between the Wellcome Centre for Human Genetics and the Big Data Institute supported by Health Data Research UK and the NIHR Oxford Biomedical Research Centre. The views expressed are those of the author(s) and not necessarily those of the NHS, the NIHR or the Department of Health. DAC was supported by the Pandemic Sciences Institute at the University of Oxford; the National Institute for Health Research (NIHR) Oxford Biomedical Research Centre (BRC); an NIHR Research Professorship; a Royal Academy of Engineering Research Chair; the Wellcome Trust funded VITAL project (grant 204904/Z/16/Z); the EPSRC (grant EP/W031744/1); and the InnoHK Hong Kong Centre for Cerebro-cardiovascular Engineering (COCHE). The ADH group at the Nuffield Department of Primary Care Health Sciences is supported by the National Institute for Health and Care Research (NIHR) Applied Research Collaboration Oxford and Thames Valley at Oxford Health NHS Foundation Trust. The views expressed are those of the author(s) and not necessarily those of the NHS, the NIHR or the Department of Health and Social Care.

## Data availability

The data reported in this paper are available via application directly to the UK Biobank, https://www.ukbiobank.ac.uk.

