## Supplemental Material for "Dysregulated Immune Proteins in Plasma in the UK Biobank Predict Multiple Myeloma 12 years Before Clinical Diagnosis"

**Plasma-based B-Cell Proteins in Healthy People Help Predict Multiple Myeloma 12 Years Before Diagnosis in the UK Biobank**
**Supplementary Materials**

#### Supplemental Method 1

**Selection of *a priori* features for baseline model**

The following features are evaluated in Constantinos et al.[1] in a UK primary care setting:

- demographics (age, sex, and BMI)
- symptoms (back, chest, bone, rib, and joint pain, shortness of breath, recurrent chest infections, fatigue, nosebleeds, bruising, fracture, weight loss, and nausea),
- blood test results (Full Blood Count components, inflammatory markers — Erythrocyte Sedimentation Rate (ESR), C-reactive Protein (CRP), and Plasma Viscosity — calcium, and creatinine)

After model development, their final two models contained the features:

- Sex
- Age
- Back pain
- Chest pain
- Rib pain
- Nosebleeds
- Haemoglobin
- White Cell Count
- Platelets
- Mean Corpuscular Volume (MCV)
- (ESR) (full model)
- (Calcium) (full model)

Given this is the most contemporary clinical myeloma risk prediction model developed in a UK population in the literature we elected to use this as our reference (baseline) model with a few modifications:

**Symptoms**

A history of rib pain and nosebleeds were not asked in UKB (https://biobank.ndph.ox.ac.uk/ukb/ukb/docs/TouchscreenQuestionsMainFinal.pdf). Given these symptoms each occurred in only 1.6% (rib pain) and 2.8% (nosebleeds) of myeloma cases in Constantinos et al. respectively, it was assessed that the impact of excluding these features on model performance would be very limited.

**Inflammatory marker**

In Constantinos et al. univariable analysis was used to select the inflammatory marker with the highest hazard ratio for inclusion in the multivariable analysis, as inflammatory marker results were found to be highly correlated. However, UKB did not do ESR on participants but has nearly complete baseline CRP data on all participants (https://biobank.ndph.ox.ac.uk/showcase/field.cgi?id=30710). Thus, CRP was included in place of ESR in our model.

**Deviations from the published model**

Constantinos et al. undertook various modifications of the data to help with model performance and convergence. Specifically, continuous variables were centred and rescaled and fractional polynomials were used to identify the optimal functional form of continuous variables: BMI, age, and blood test results.

We opted not to do this step for the following reasons:

1- Ours is a baseline benchmark model to compare the proteomics model to and we wished to keep coefficients largely interpretable so that they could be better understood
2- Schoenfeld residuals indicated that none of these features ultimately included in the Cox model violated the proportional hazards assumption in their native form meaning their functional form did not need to be changed for this reason

#### Supplemental Method 2

**Study design and participants**

The UK Biobank (UKB) is a population-based prospective cohort study comprising approximately half a million participants aged 40-69 years, recruited between 2006 and 2010 from 22 assessment centres across England, Wales, and Scotland.[1] Baseline data were collected through questionnaires, verbal interviews with trained nurses, physical examinations, and biological samples.[2] Follow-up information is obtained through linked electronic medical records, including death and cancer registries and hospital inpatient records. The study has ethical approval from the National Health Service Northwest Multicentre Research Ethics Committee (06/MRE08/65), and all participants provided written informed consent.

**Plasma Proteomics data**

The UKB Pharma Proteomics Project (UKB-PPP) involved the collection and processing of plasma samples from over 54,000 participants to measure the relative abundance of 2,923 unique proteins using the Olink Proximity Extension Assay (PEA) where the relative concentrations of the proteins are ascertained via next-generation sequencing of DNA tags attached to protein-specific antibodies.[3] Samples were selected through a combination of pre-selection by consortium members and stratified random sampling based on age, sex, and recruitment centre. Protein levels were expressed as Normalised Protein eXpression (NPX) values, with data normalisation steps applied to account for plate-to-plate and batch-to-batch variations. Initial quality control (QC) measures implemented by Olink included the use of internal controls and duplicate samples to monitor intra- and inter-plate variability. Additional comprehensive QC procedures performed by the UKB-PPP included removing outliers identified through principal component analysis, removing data with assay warnings, and identifying and removing likely swapped samples. These have been described in detail previously[3] and were implemented prior to the release of data on UKB showcase. Within the study, three proteins were removed prior to analysis for having more than 30% missing data.

**Inclusion and Outcome Assessment**

This study was restricted to participants with baseline plasma proteomics data, without prevalent myeloma at baseline. Outcome data were ascertained through linkage to national electronic medical records of death and cancer registries and hospital inpatient records. Myeloma diagnoses were initially identified using cancer registry data, with cutoff dates of December 31, 2020, for England, November 30, 2021, for Scotland, and December 31, 2016, for Wales. This primary source was used to identify both prevalent and incident cases based on ICD codes. To ensure comprehensive outcome identification, we used hospital inpatient records to further identify incident myeloma cases, with data available up to October 31, 2022, for England, August 31, 2022, for Scotland, and May 31, 2022, for Wales. Censoring for non-cases was at either date of death or end of hospital data linkage, whichever occurred first. The primary endpoint was the first incident occurrence of ICD-10 code C90.0. Person-years of follow-up were calculated from recruitment until the first myeloma registration, cancer-related death, death from other causes, loss to follow-up, or hospital censoring date.

**Analysis pipeline**

The pipeline for ML-based feature selection was adapted from an approach described by Lui et al.[4] In this approach, a tree-based eXtreme Gradient Boosting (XGBoost) machine learning algorithm is applied to a large tabular dataset to identify potentially novel features associated with the outcome of interest. Our pipeline consists of several key steps: first, all possible proteomic predictors (n=2920) were included in an XGBoost model, and SHAP values used to identify the top 10 most important predictors to this model. These top 10 features were then utilised in two ways: first, to create a proteomics-only Cox model, and second, by combining them with *a priori* clinical predictors of myeloma diagnosis. As a result, three Cox models were developed: a proteomics Cox model, a combined Cox model, and a clinical Cox model that included only clinical predictors. The dataset was divided into training and test datasets at a 4:1 ratio, with all model development, and tuning conducted on the training dataset. The performance of these models was evaluated on the test dataset using time-dependent area under the receiver-operating-characteristic curve (AUC) and concordance indexes (C-indexes).

**Feature selection**

Feature selection is required for any analytical model and can be achieved through various methods, each with their own trade-offs. Tree-based XGBoost is an ensemble learning method that sequentially constructs decision trees, each minimising the residuals of its predecessors, with the final prediction being the weighted sum of these trees' outputs.[5] This technique efficiently reveals non-linear relationships among correlated features in large datasets. We chose to use model-based feature selection using a tree-based XGBoost model because of its ability to rank numerous features, handle missing data, and take account of non-linear relationships between features and outcomes. The ability to capture interactions between features is also of interest in understanding the relationship between 2920 plasma proteins many of which are yet to be fully understood. In addition, this approach enabled us to identify candidate features with non-linear methods and use classical models for final evaluation of associations.

The model was trained on a "Cox loss" function, by trying to minimise the negative log partial likelihood of the Cox model. XGBoost handles missing data intrinsically by learning a decision that optimises the loss function for each feature with missing data. Thus, during training data are considered missing not at random. Hyperparameters were tuned via grid search with five-fold cross validation. The optimal hyperparameters estimated were column sample by tree, $\phi$ = 0.8; $\gamma$ = 0; learning rate, $\eta$ = 0.1; maximum depth, $d_{max}$ = 7; minimum child weight, $\omega_{min}$ = 2; number of trees, $T$ = 500; alpha regularisation, $\alpha$ = 0, lambda regularisation, $\lambda$ = 4; and subsample, $\rho$ = 0.6.

SHAP values are based on cooperative game theory and provide a unified measure of feature importance by assigning each feature an importance value for a particular prediction.[6] SHAP values are particularly useful for evaluating feature importance in our tree-based XGBoost model because they offer consistent and interpretable insights into the contribution of each feature to the prediction. This is helpful given the potential complexity of the feature space. We present two types of SHAP plots: the mean absolute SHAP value plot and the bee-swarm plot. The mean absolute SHAP value, defined mathematically as $\frac{1}{n}\sum_{i=1}^{n} | S_{i,j} |$, where $S_{i,j}$ represents the SHAP value of feature *j* for instance $i$, and $n$ is the total number of instances, for the top 10 features are plotted to show the average absolute impact of each feature on the model's predictions, highlighting the overall importance of each feature. In contrast, the bee-swarm plot provides a more granular view by showing the distribution of SHAP values for each feature across all samples, thus illustrating the variability and the direction of the feature's impact. From these plots, we identified the top 10 features with the highest mean absolute SHAP values for subsequent evaluation.

**Statistical analysis**

For the statistical analysis we first constructed a "clinical" model based on covariates chosen *a priori* from a UK population-based study (see Supplementary Info 1 for details).[7] We then augmented this model with the top 10 SHAP features and finally compared it to one with only the proteomic features. Before inclusion in models, we first assessed pairwise correlations of these features and then examined the highly correlated features as this would preclude the inclusion of both colinear features. Where a pair of features was identified as highly correlated (> 0.9), we removed the feature with most missing data. Pairwise corelation tests were performed with Spearman's rank coefficient for pairs of numeric features, Point-Biserial correlations for all numeric features with the binary features, and a Cramér's V correlation heatmap for the three binary features (sex, back pain, and chest pain).

Multiple imputation was performed on all numerical features prior to Cox models. Ten rounds of imputation were done on the training set using the *IterativeImputer* package from scikit-learn under the assumption of the data being missing at random. For the features "chest pain" and "back pain", any response other than yes was presumed to be no. Due to very low variance on its measured scale, Calcium was normalised using z-transform for fitting of the Cox models.

To compare model performance, Harrel's C-index and time-varying receiver operating characteristic (ROC) curves were calculated at years 4, 8, 12, and 16 (n cases in test set = 8, 11, 7, 1) to capture the changes in discrimination ability of the models. We chose these time intervals because they allow us to evaluate the models at comparable intervals. Unlike a standard ROC curve, which evaluates the model's performance at a single time point, a time-varying ROC curve assesses the model's predictive performance across multiple time points, reflecting how well it distinguishes those who will experience the event at various stages of follow-up. This approach allows for a better evaluation of the model's ability to predict the timing of events, accommodate the dynamic nature of risk, while handling censored data appropriately. This approach is especially useful in the context of reverse causality as myeloma diagnoses are often delayed.

The proportional hazard assumption of Cox models was assessed using scaled Schoenfeld residuals. Collinearity was assessed by computing the variance inflation factor; values less than ten were considered acceptable. Statistical tests were two-tailed and performed using a 5% significance level. Unless otherwise specified, normally distributed variables were compared using t-tests, non-normally distributed variables with Wilcoxon rank-sum tests, and categorical variables with chi-squared tests.

**Sensitivity analyses**
To evaluate the temporal relationship between features and time to diagnosis we evaluated a time-stratified modelling approach, fitting separate Cox models for cases diagnosed within the first five years of follow-up and another one for those beyond five years. In addition, to evaluate the impact of known baseline monoclonal gammopathy of unknown significance (MGUS), we fitted Cox models on the same training data excluding all participants with prevalent and incident MGUS (defined by a diagnostic ICD10 code of D47.2 in summary hospital statistics data).

**Software**
For machine learning and data imputation, scikit-learn version 1.5, XGBoost version 2.0, and SHAP version 0.45 were implemented in Python version 3.9 in a virtual environment. R version 4.4.2 was used for hypothesis testing, summary statistics, and survival analyses.

#### Supplemental Table 1

**Baseline cohort characteristics by myeloma outcome status.** Mean (standard deviation) are presented for continuous variables except time (median, [interquartile range]), frequency (percentage) are reported for categorical variables.

| Variable | | Without incident myeloma (n=52825) | With incident myeloma (n=174) | N |
|---|---|---|---|---|
| **Time, years** | 13.6 [12.9, 14.3] | 13.62 [12.9, 14.3] | 7.2 [4.1, 10.2] | 52999 |
| **Age, years** | 57.3 (8.2) | 57.3 (8.2) | 61.1 (6.7) | 52999 |
| **Sex, male** | 24425 (46.1) | 24331 (46.1%) | 94 (54.0%) | 52999 |
| **Back pain, yes** | 13978 (26.4) | 13920 (26.4%) | 58 (33.3%) | 52999 |
| **Chest pain, yes** | 8949 (16.9) | 8910 (16.9%) | 39 (22.4%) | 52999 |
| **Haemoglobin, g/dL** | 14.15 (1.26) | 14.1 (1.3) | 14.1 (1.2) | 51244 |
| **MCV, fL** | 91.11 (4.65) | 91.1 (4.7) | 91.9 (4.8) | 51244 |
| **Platelet count, x10^11 cells/Litre** | 252.70 (60.10) | 252.7 (60.1) | 253.6 (71.9) | 51244 |
| **White blood cell count, x10^11 cells/Litre** | 6.89 (2.12) | 6.9 (2.1) | 6.7 (1.8) | 51243 |
| **CRP, mg/L** | 2.66 (4.48) | 2.7 (4.5) | 3.2 (6.3) | 50271 |
| **Calcium, mmol/L** | 2.38 (0.09) | 2.4 (0.1) | 2.4 (0.1) | 46194 |

***MCV***: *Mean Corpuscular Volume.* ***CRP***: *serum C-Reactive Protein.* ***Calcium*** *serum calcium.*

#### Supplemental Table 2

**Summary of biological information on the top 10 proteins.** A summary of protein function information derived from uniprot[8], location and single cell expression from the Human Protein Atlas[9], and therapeutic information from the open targets platform[10] for the top 10 identified proteins.

| Protein | Function | Location | Expression | Therapeutic Target in Myeloma |
|---|---|---|---|---|
| **SLAMF7** | Immune cell signalling | Membrane, Intracellular | Enhanced: Plasma cells, Dendritic cells, Monocytes, Schwann cells, NK-cells | Yes, SLAMF7 binding antibody elotuzumab, authorised for use by the EMA |
| **TNFRSF17 (BCMA)** | Receptor for BAFF & APRIL, mediates B cell maturation and promotes survival | Membrane, Intracellular | Enriched: Plasma cells | Yes, BCMA binding antibodies, Elranatamab, Teclistamab authorised for use by the EMA |
| **QPCT** | Enzyme for the biosynthesis of pyroglutamyl peptides | Secreted, Intracellular | Enhanced: Melanocytes, Langerhans cells, Pancreatic endocrine cells, Enteroendocrine cells | No |
| **LY9 (SLAMF3)** | Immune cell signalling | Secreted, Membrane | Enhanced: B-cells, Plasma cells, Dendritic cells, T-cells, Langerhans cells | No |
| **SLAMF1** | Immune cell signalling | Secreted, Membrane | Enriched: Plasma cells, T-cells, B-cells | No |
| **CNTN5** | Mediates cell surface interactions during nervous system development | Intracellular | Enriched: Inhibitory neurons, Excitatory neurons | No |
| **TNFRSF13B (TACI)** | Receptor for APRIL and BAFF; involved in the stimulation of B- and T-cell function | Membrane, Intracellular | Enriched: Plasma cells, B-cells | No |
| **TNFSF13 (APRIL)** | Cytokine that binds to TACI and BCMA; regulates tumour cell growth | Secreted, Intracellular | Enhanced: Mesothelial cells, Macrophages, Schwann cells, Basal squamous epithelial cells, Erythroid cells | No |
| **TNFSF13B (BAFF)** | Cytokine that binds to TACI and BCMA; binds to the same 2 receptors as APRIL. | Secreted, Membrane, Intracellular | Enhanced: Monocytes, Langerhans cells, Hofbauer cells, Macrophages, Kupffer cells | Tabalumab, a BAFF inhibiting antibody in previous phase I and II trials. |
| **TIMP1** | Metalloproteinase inhibitor. Also functions as a growth factor. | Secreted, Intracellular | Enhanced: Mesothelial cells, Monocytes, Langerhans cells, Fibroblasts, Sertoli cells, Pancreatic endocrine cells | No |

***SLAMF7**: Signalling Lymphocytic Activation Molecule Family Member 7. **TNFRSF17**: Tumour Necrosis Factor Receptor Superfamily Member 17. **BCMA**: B-cell Maturation Antigen. **QPCT**: Glutaminyl-Peptide Cyclotransferase. **LY9**: Lymphocyte Antigen 9. **SLAMF3**: Signalling Lymphocytic Activation Molecule Family Member 3. **SLAMF1**: Signalling Lymphocytic Activation Molecule Family Member 1. **CNTN5**: Contactin 5. **TNFRSF13B**: Tumour Necrosis Factor Receptor Superfamily Member 13B. **TACI**: Transmembrane Activator and CAML Interactor. **CAML**: Calcium-Modulating Cyclophilin Ligand; **TNFSF13**: Tumour Necrosis Factor Superfamily Member 13. **APRIL**: A Proliferation-Inducing Ligand. **TNFSF13B**: Tumour Necrosis Factor Superfamily Member 13B. **BAFF**: B-cell Activating Factor. **TIMP1**: Tissue Inhibitor of Metalloproteinases 1. **EMA**: European Medicines Agency.*

#### Supplemental Figure 1

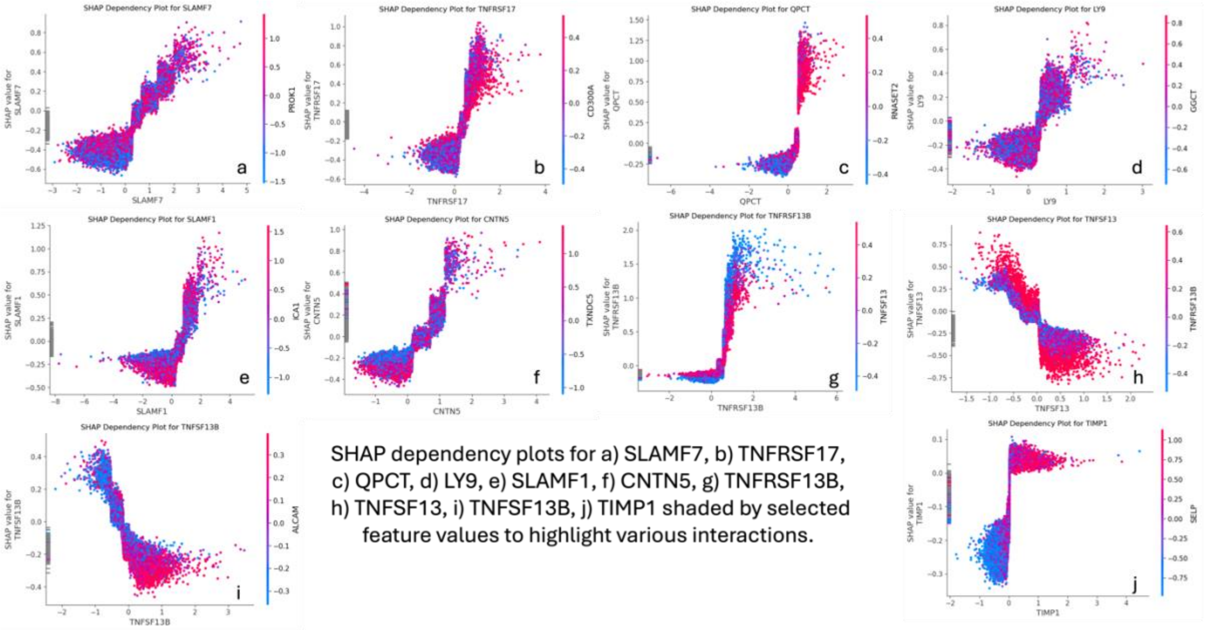

**Supplemental figure 1.1:** SHAP Dependency plots

The figure presents ten SHAP dependency plots (a-j), each depicting the relationship between the values of one of the top 10 most important features from the first (feature selection) XGBoost model and their corresponding SHAP values. These SHAP values represent the feature's contribution to the model's prediction for incident myeloma. Each plot is shaded based on the values of another feature to highlight potential interactions between features. The purpose of these plots is to illustrate both the direct effect of each feature on the model output and any interaction or non-linear relationship with other features.

In the plots:

x-axis - shows the value of the feature for each observation.

y-axis - shows the SHAP value (impact on model output).

Colour shading - represents the values of a second feature, automatically chosen by SHAP to highlight interaction effects.

**Interactions:**

Several interactions are visible based on the shading in different plots. Important interactions include:

1. **TNFRSF17 (BCMA) (Panel b):**
    - **Interaction:** The red shading (indicating high values of CD300A) is concentrated in the upper right of the plot, suggesting that high values of TNFRSF17 (BCMA) in combination with high values of the interacting feature led to a stronger positive SHAP contribution. This indicates a potential synergistic effect between TNFRSF17 (BCMA) and CD300A.
2. **TNFRSF13B (TACI) (Panel g):**
    - **Interaction:** The plot shows a positive dependency on TNFRSF13B (TACI), and the shading emphasises that the SHAP values increase much more rapidly for high values of both TNFRSF13B and the interacting feature

(TNFSF13/APRIL) seen by the red points clustering on the right-hand side). This strong interaction suggests that these two features together lead to a higher model contribution.

3. **TNFSF13 (APRIL) (Panel h):**
   - **Interaction**: This is different representation of the interaction between TNFRSF13B (TACI) and TNFSF13 (APRIL). Specifically, when TNFSF13 values are low (around -1.5 to 0 on the x-axis), the SHAP value can be positive or close to zero, but the effect depends heavily on the value of TNFRSF13B (the colour shading). For example, when TNFRSF13B is low (blue shading), the SHAP value of TNFSF13 tends to be lower or even negative, whereas at higher TNFRSF13B values (red shading), the SHAP values are much more likely to be positive. Similarly, at higher values of TNFSF13 (x-axis values around 0 to 2), the SHAP values are more likely to be negative, and the interaction with TNFRS13B changes. Here, higher TNFRSF13B values (red-shaded points) lead to much stronger negative SHAP values compared to lower TNFRSF13B values (blue-shaded points). In essence, the interaction means that high values of TNFRSF13B increase the magnitude of the SHAP value at all values of TNFSF13

**Non-Linear Associations:**
Non-linear patterns are present in several plots, indicating that the effect of a feature on the model output changes at different feature values. Important non-linear associations include:

1. **QPCT (Panel c):**
   - **Non-Linearity:** The association between QPCT and its SHAP values is distinctly non-linear. For lower values of QPCT, there is little impact on the model output, but after a threshold (around 1.5), the SHAP values rapidly increase. This non-linear behaviour indicates that QPCT only becomes influential above a certain value.
2. **SLAMF1 (Panel e):**
   - **Non-Linearity:** The SHAP values for SLAMF1 are low negatives for all negative values of SLAMF1, rise sharply with increasing SLAMF1 values between 0 and 2 and then reaches a saturation point above values of 2, beyond which further increases in SLAMF1 result in a minimal change in the SHAP value.
3. **TNFRSF13B (Panel g):**
   - **Non-Linearity:** TNFRSF13B shows a strong non-linear relationship with SHAP values, with a sharp rise in model impact only after TNFRSF13B reaches around 0 on the x-axis. This steep rise indicates that TNFRSF13B becomes important only at higher values.

In summary, this figure highlights key interactions between proteomic features and strong non-linear effects, revealing how these top predictors contribute to the XGBoost model for myeloma prediction.

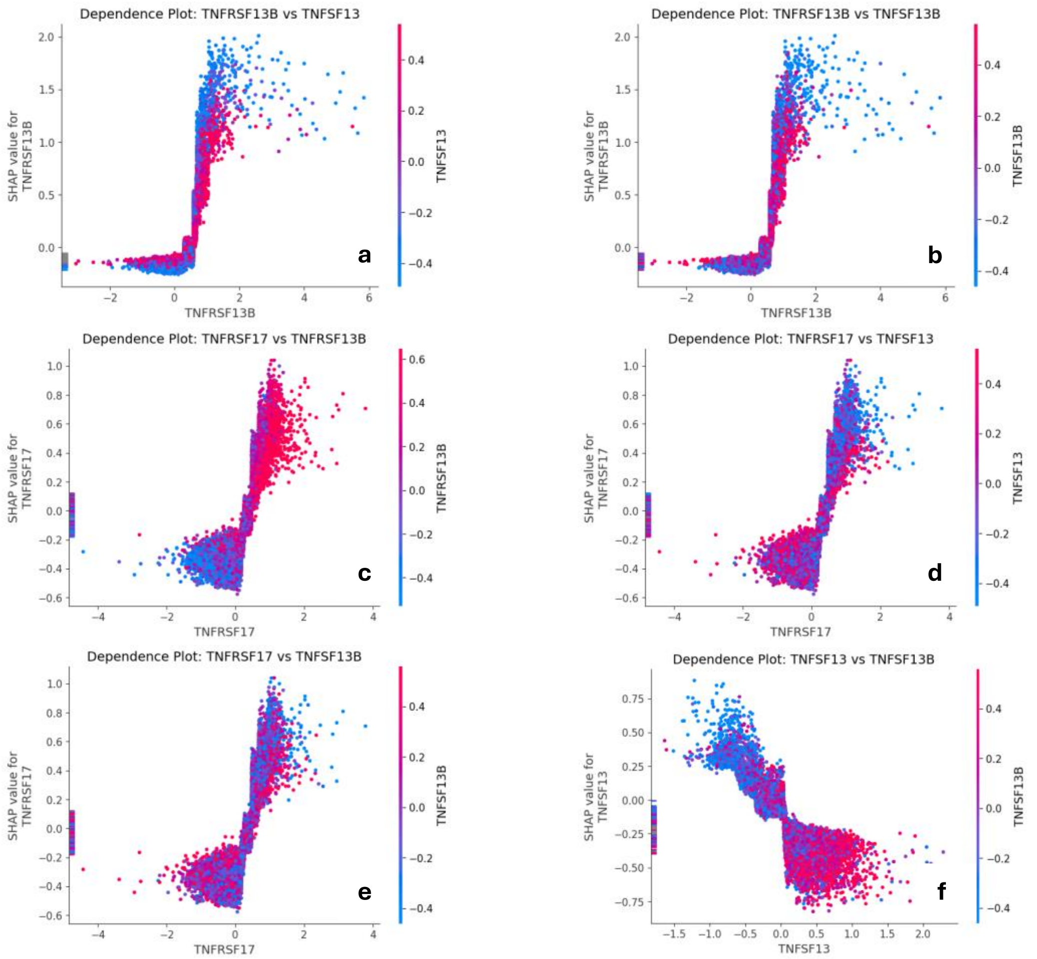

**Supplemental figure 1.2:** SHAP Dependency plots specifically focusing on the interacting receptor ligand combinations of TNFRSF17 (BCMA), TNFRSF13B (TACI), TNFSF13, (APRIL), and TNFSF13B (BAFF) with **a** presenting TACI and APRIL, **b** TACI and BAFF, **c** BCMA and TACI, **d** BCMA and APRIL, **e** BCMA and BAFF, and **f** APRIL and BAFF. As discussed above, there are noticeable interactions between TACI and APRIL (**a**), with the one between TACI and BAFF (**b**) much weaker as seen by the less clear delineation and separation of the red and blue dots at either end of the values especially at low levels of TACI.

#### Supplemental Figure 2

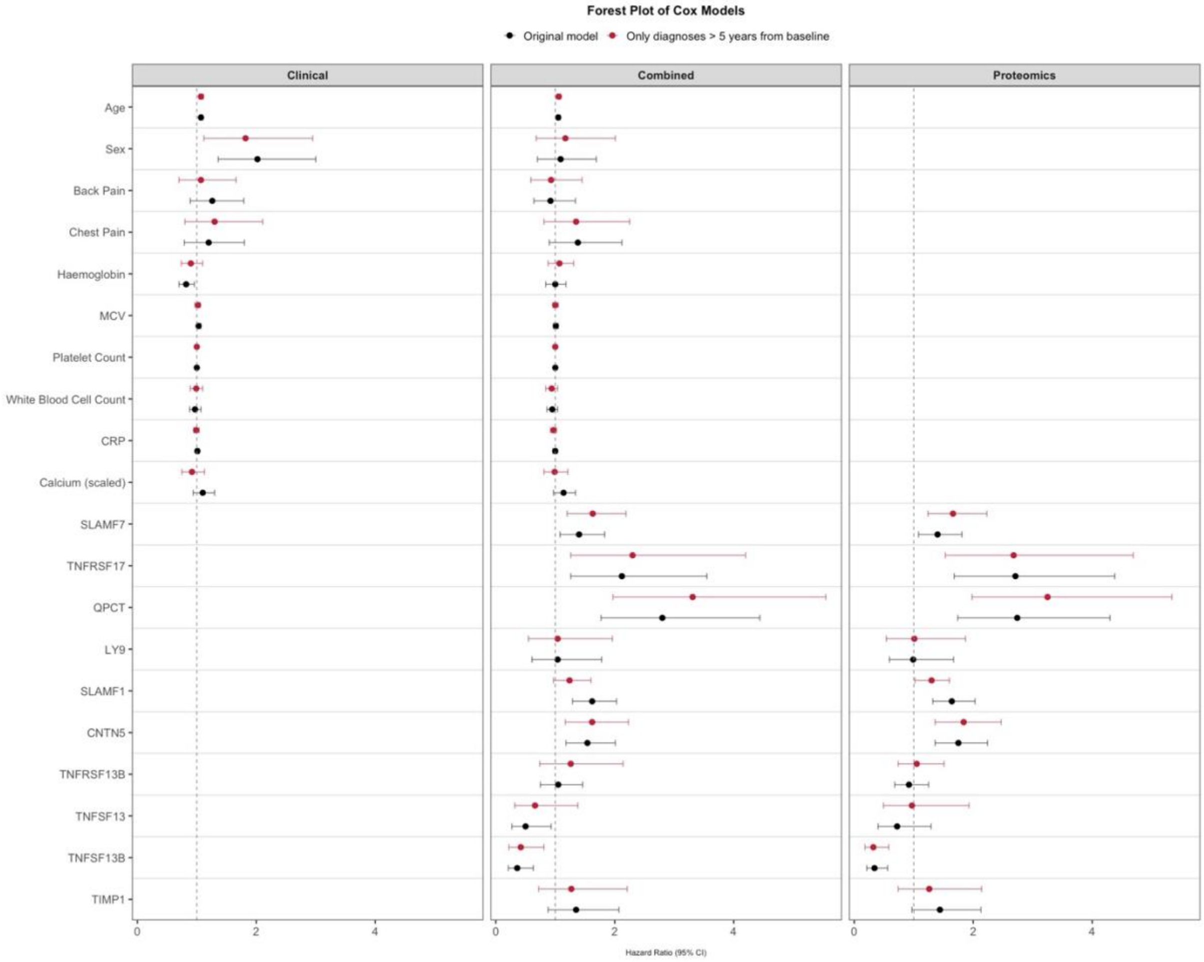

**Supplemental figure 2:** Forest plot comparing original Cox model estimates (black) with a sensitivity analysis excluding all participants diagnosed with MM within 5 years of baseline (red)

This figure presents three forest plots comparing hazard ratios (HR) and 95% confidence intervals (CI) from Cox proportional hazards models based on the clinical features (left panel), proteomics markers (right panel), and the combined features (centre) for predicting myeloma diagnosis across two different time intervals: >5 years from baseline (red), and all incident diagnoses (black). Dotted lines at HR = 1 indicate no association.

### Supplemental Figure 3

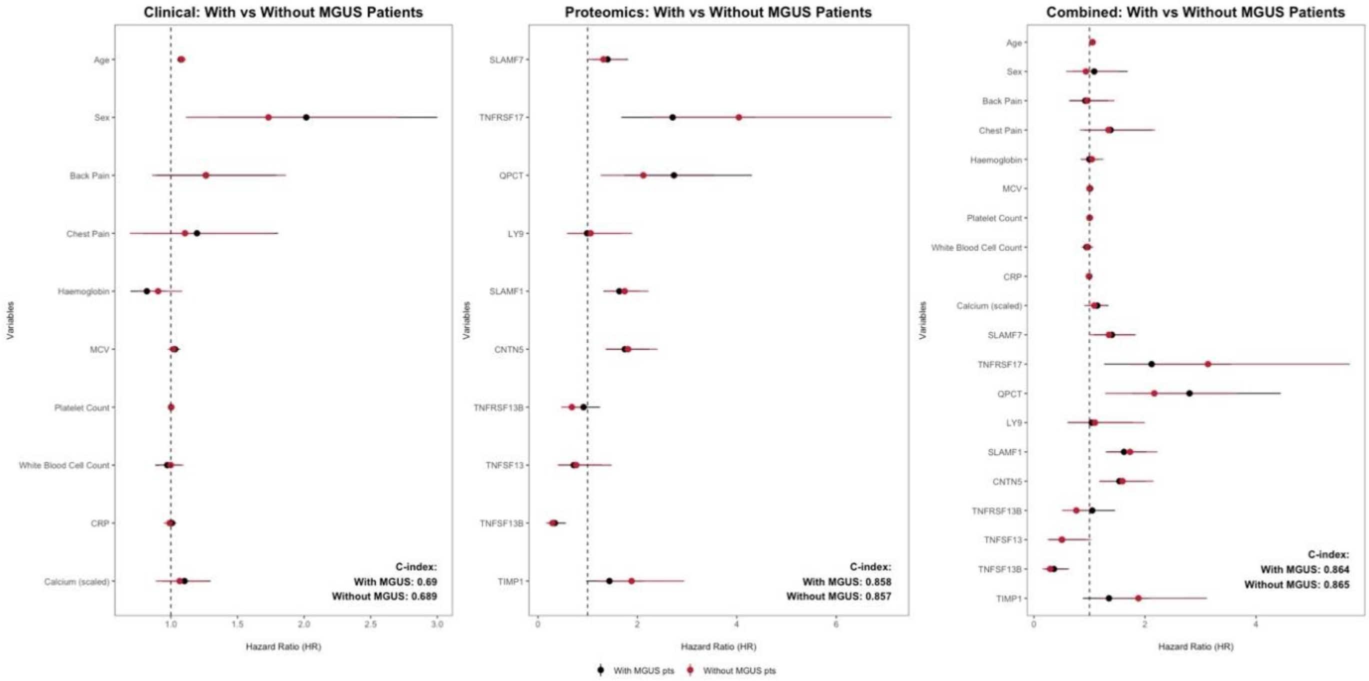

**Supplemental figure 3:** Forest plot comparing original Cox model estimates (black) with a sensitivity analysis excluding all participants with MGUS diagnoses (red).

There are 8 cases of prevalent MGUS identified in hospital record data, 4 of whom subsequently developed incident myeloma and there are 155 cases of incident MGUS identified in hospital record data, 20 of whom also developed incident myeloma. A sensitivity analysis excluding these participants from the analysis is shown in the above figure in red ("Without MGUS pts"). While the magnitudes of certain proteomic associations (e.g. TNFRSF17 and QPCT) change, none of the directions of association change and many associations retain their statistical significance (95% confidence intervals demonstrated by the associated whiskers on the forest plot). Performance of the various models, as assessed via c-index on the training dataset, remains unchanged. Dotted lines at HR = 1 indicate no association.
